# A Nonlinear Observer to Estimate the Effective Reproduction Number of Infectious Diseases

**DOI:** 10.1101/2021.03.02.21252730

**Authors:** Agus Hasan

## Abstract

In this paper, we design a Nonlinear Observer (NLO) to estimate the effective reproduction number (*ℛ*_*t*_) of infectious diseases. The NLO is designed from a discrete-time augmented Susceptible-Infectious-Removed (SIR) model. The observer gain is obtained by solving a Linear Matrix Inequality (LMI). The method is used to estimate *ℛ*_*t*_ in Jakarta using epidemiological data during COVID-19 pandemic. If the observer gain is tuned properly, this approach produces similar result compared to existing approach such as Extended Kalman filter (EKF).

## I. Introduction

Governments around the world are using the reproduction numbers as criteria when deciding public health policies during COVID-19 pandemic. In principal, there are two types of reproduction numbers: basic reproduction number and effective reproduction number. The basic reproduction number (denoted by *ℛ*_0_) shows the average expected number of cases generated by one case in a population where all individuals are susceptible. The effective reproduction number (denoted by *ℛ*_*t*_) shows the average expected number of cases generated in the current state of a population. In practice, *ℛ*_0_ is used to determine how many population that needs to be immune to reach herd immunity. In this case, the herd-immunity threshold is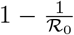. On the other hand, *ℛ*_*t*_ is used to monitor transmissions of the disease in a population during the outbreak. Hence, this number is usually used as one of the main criteria to evaluate the public health policies. Theoretically, *ℛ*_*t*_ needs to be below 1 to ensure the transmission is under control.

Many researchers have provided calculations to estimate *ℛ*_*t*_ using different approaches, e.g., Bayesian estimation [1], serial interval [2], Extended Kalman filter [3], and parameter fitting [4]. If the estimation parameters are tuned properly, all of these approaches will provide similar pattern with small variation. Once the estimated *ℛ*_*t*_ is obtained, we can create short-term forecasts to determine different reopening scenarios [5]–[7].

The aim of this paper is to provide a novel approach to estimate the effective reproduction number *ℛ*_*t*_of infectious diseases. To this end, we design a Nonlinear Observer (NLO) from a discrete-time augmented Susceptible-Infectious-Removed (SIR) model. The method is efficient in the sense that the epidemiological data is injected directly into the model once we found a constant observer gain. The observer gain is obtained by solving a Linear Matrix Inequality (LMI). While the majority of estimation methods are based on stochastic process, this new approach is deterministic. The Confidence Interval (CI) provided in our estimation is inherited from the uncertainty in the infectious time and not from the method itself.

The paper is organized as follow. In Section II, we derive a discrete-time augmented SIR model. In this section, we consider the time-varying *ℛ*_*t*_ as an augmented state. Furthermore, we assume its value is a piece-wise constant function. In Section III, we derive a sufficient condition for the observer gain in terms of LMI. Simulation results using epidemiological data from Jakarta is presented in Section IV. Finally, conclusions are given in Section V.

## II. Model

We use a simple SIR model in this paper for several reasons. First, it can be used to described transmissions of many infectious diseases. Furthermore and most importantly, the actual data of the three compartments are available in most outbreak events. The removed compartment consists of individuals who are either recovered or died. Assuming constancy of population *N*, the SIR model can be written as follow [8]:

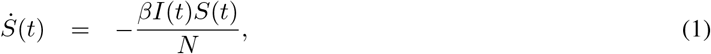

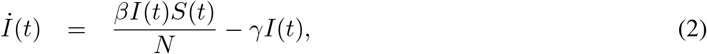

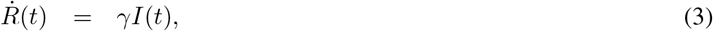

where *S* denotes the number of susceptible individuals, *I* denotes the number of infected individuals or active cases, and *R* denotes the number removed individuals. The model has two parameters: the transmission rate *β* and the removal rate *γ*. By definition, *β* is the average number of contacts per person per time, multiplied by the probability of disease transmission in a contact between a susceptible and an infectious individual. Thus, in principal *β* is time-varying due to interventions. For this reason, in the remaining of this paper we consider *β* as a parameter that depends on time *t* and is unknown. On the other hand, the removal rate *γ* is an inverse of the average infectious time, i.e., 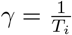 The infectious time can be obtained through medical data and is usually known together with its Confidence Interval (CI). Taking into account reduction in the number of susceptible individuals, the effective reproduction number can be estimated as follow [9]:

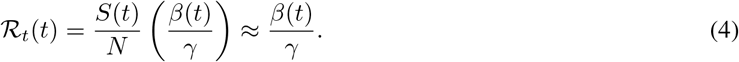

Discretizing (1)-(3) using the Euler discretization method, substituting *β*(*t*) = *γ ℛ*_*t*_(*t*) into the model, and augmenting *ℛ*_*t*_(*t*) as a new state variable, we obtain the following discrete-time augmented SIR model:

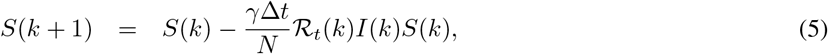

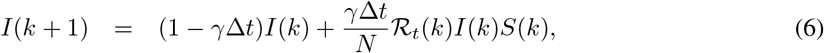

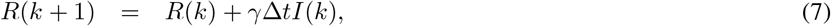

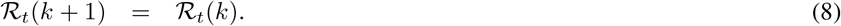

Remark that, in the last equation we assume *ℛ*_*t*_ as a piece-wise constant function with jumps every time new data come in. To simplify the model, let us define:

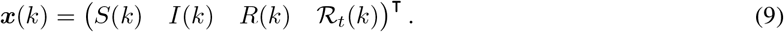

The discrete-time augmented SIR model (5)-(8), can be written as the following nonlinear state-space representation:

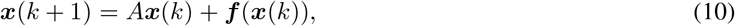

Where

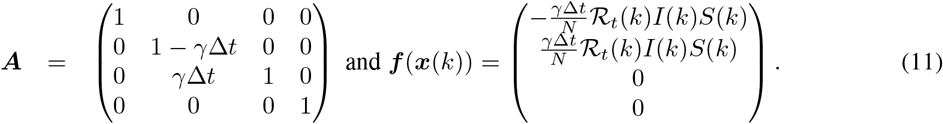

Since data for *S*(*k*), *I*(*k*), and *R*(*k*) are available, the measurement vector ***y***(*k*) ∈ ℝ^3^ is given by:

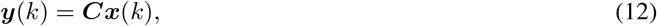

Where

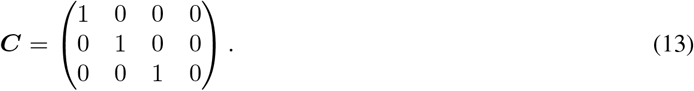

## III. Method

We design the NLO as follow:

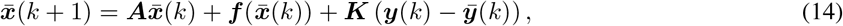

where 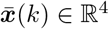 is the estimated state from the NLO and ***K***∈ ℝ^4×3^ is the observer gain to be determined later. Let 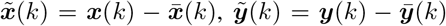 and 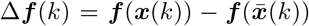. Subtracting (10) with (14), we obtain:

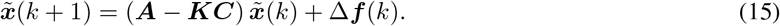

The problem is to find ***K*** such that the error 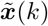 is asymptotically converges toward zero, which guarantees the estimated states converge to the actual states. Designing a NLO for a nonlinear system is not trivial and sometime impossible. Thus, we can simplify the problem using some assumptions, for example by assuming the non-linearity is locally Lipschitz and bounded. These assumptions are common when designing NLO for nonlinear systems, e.g., see [10], [11]. Therefore, the following assumptions are used in this paper:

### Assumption 1.

*The nonlinear function f is a one-sided Lipschitz, i*.*e*., *it satisfies*

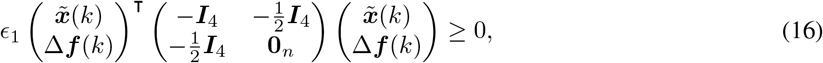

*for ϵ*_1_ > 0, *where* ***I***_4_ *denotes a* 4 × 4 *identity matrix*.

### Assumption 2.

*The nonlinear function* ***f*** *satisfies the quadratic inner-boundedness condition, i*.*e*.,

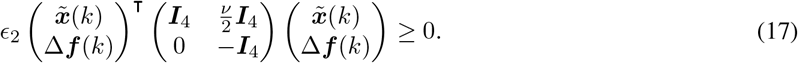

*for ϵ*_2_ > 0 *and ν* ∈ ℝ.

Utilizing the above assumptions, we can derive a sufficient condition for the observer gain in terms of LMI.

### Theorem III.1.

*Under* ***Assumption 1*** *and* ***Assumption 2***, *the error dynamics* (15) *is asymptotically stable if there exist matrices* ***G*** = ***G***^T^ *>* 0 *and* ***R***∈ ℝ^3×4^ *such that the following LMI holds*

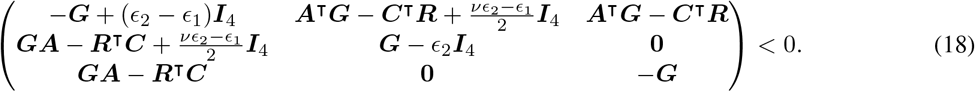

*Furthermore, the observer gain is given by:*

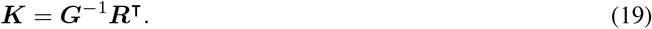

*Proof*. Let us define a Lyapunov function:

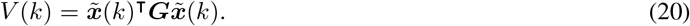

Thus, if Δ*V* (*k* + 1) = *V* (*k* + 1) − *V* (*k*), then we have:

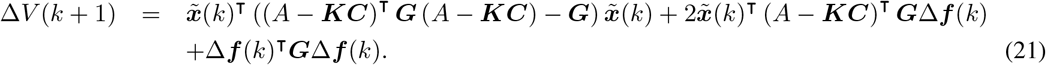

Expressing the right hand side of (21) as a matrix multiplication, we have:

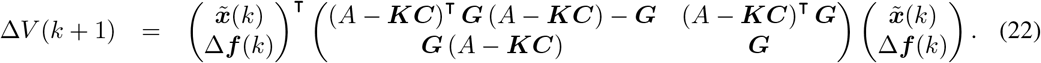

Adding the left hand side of (16) and (17) into (22), we have:

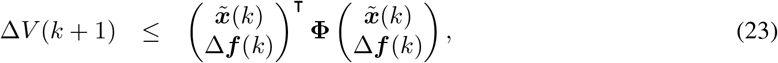

where

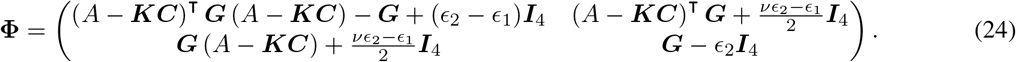

Substituting (19) and applying Schur complement to (24), then **Φ** < 0 is equivalent to (18). This completes the proof. □

## IV. Estimation of *ℛ*_*t*_ in Jakarta

Daily epidemiological data of COVID-19, such as the number of active case and the number of removed case between April 2020 until August 2020, are used in our estimation. Code and data are available in: https://github.com/agusisma/COVIDNLO. In this simulation, we use the following parameters: *ϵ*_1_ = 10, *ϵ*_2_ = 1, and *ν* = 9. Furthermore, the symmetric matrix ***G*** is chosen as ***G*** = 0.1***I***_4_, while the matrix ***R*** is chosen as:

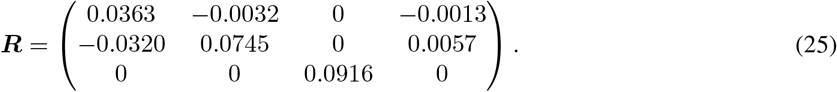

We assume the average infectious time 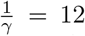 days with standard deviation of 3 days. Using these parameters, the LMI (18) is negative definite with the largest and smallest Eigenvalues are −0.0989 and −9.1294, respectively. The observer gain is then given by:

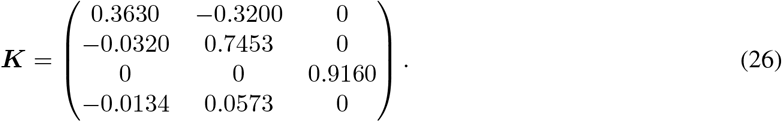

Figure 1 shows estimation results from the NLO for the daily number of active and removed case. It can be observed from the estimation errors that the NLO estimates these numbers reasonably accurate. The NLO is compared with the EKF method presented in [3] and the results can be seen from Figure 2. It can be observed that the estimation results are virtually almost identical. We should note, however, that these results are obtained after a lengthy process of trial and error when determining the matrix ***R***. Unfortunately, there is no method that can be used to determine matrix ***R*** automatically. Having said that, the main advantage of using NLO is its stability and efficiency compare to EKF, since the NLO doesn’t require calculation of inverse matrices.

**Fig. 1:**
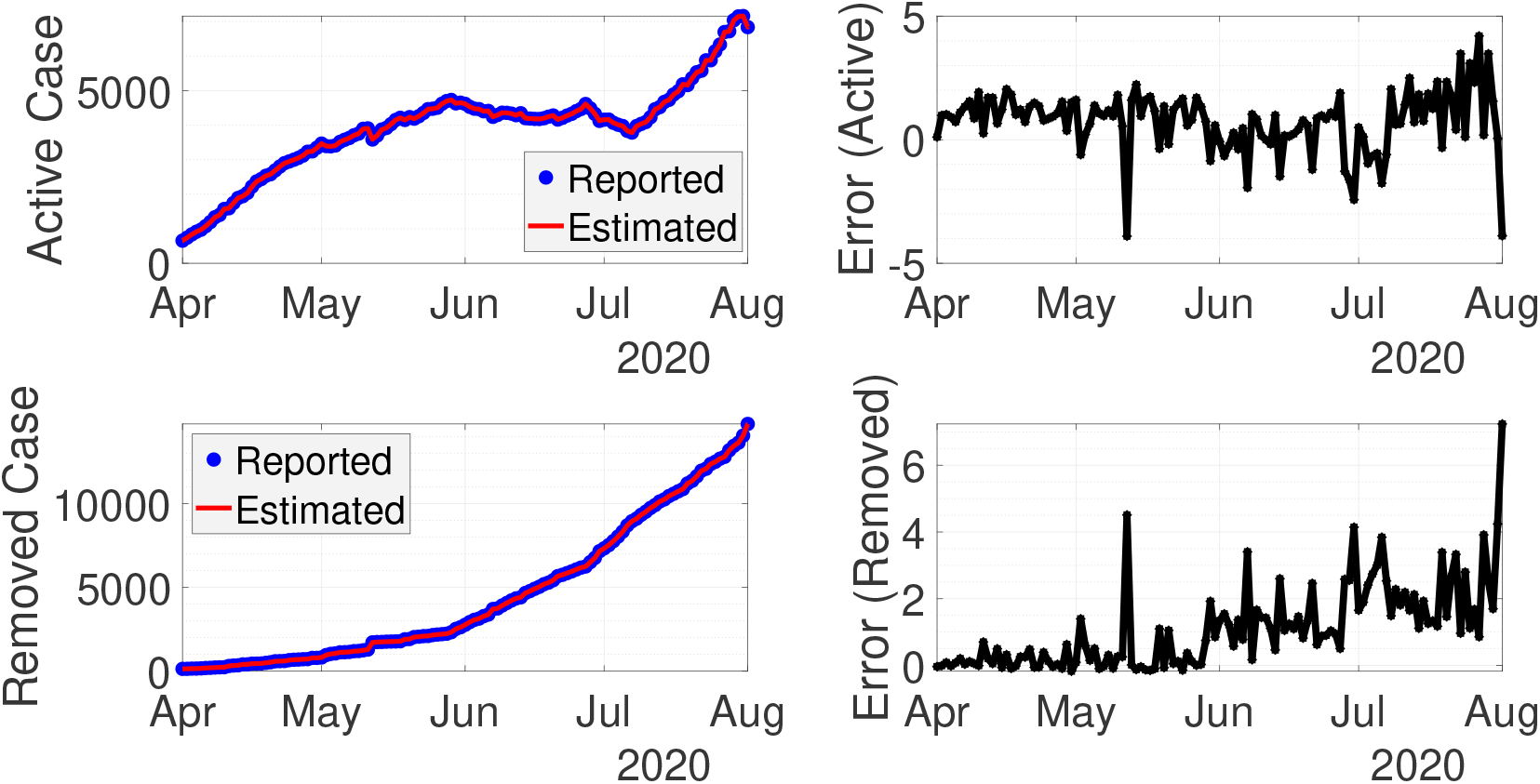
Real-time data fitting from the NLO for active and removed case with their estimation errors.

**Fig. 2:**
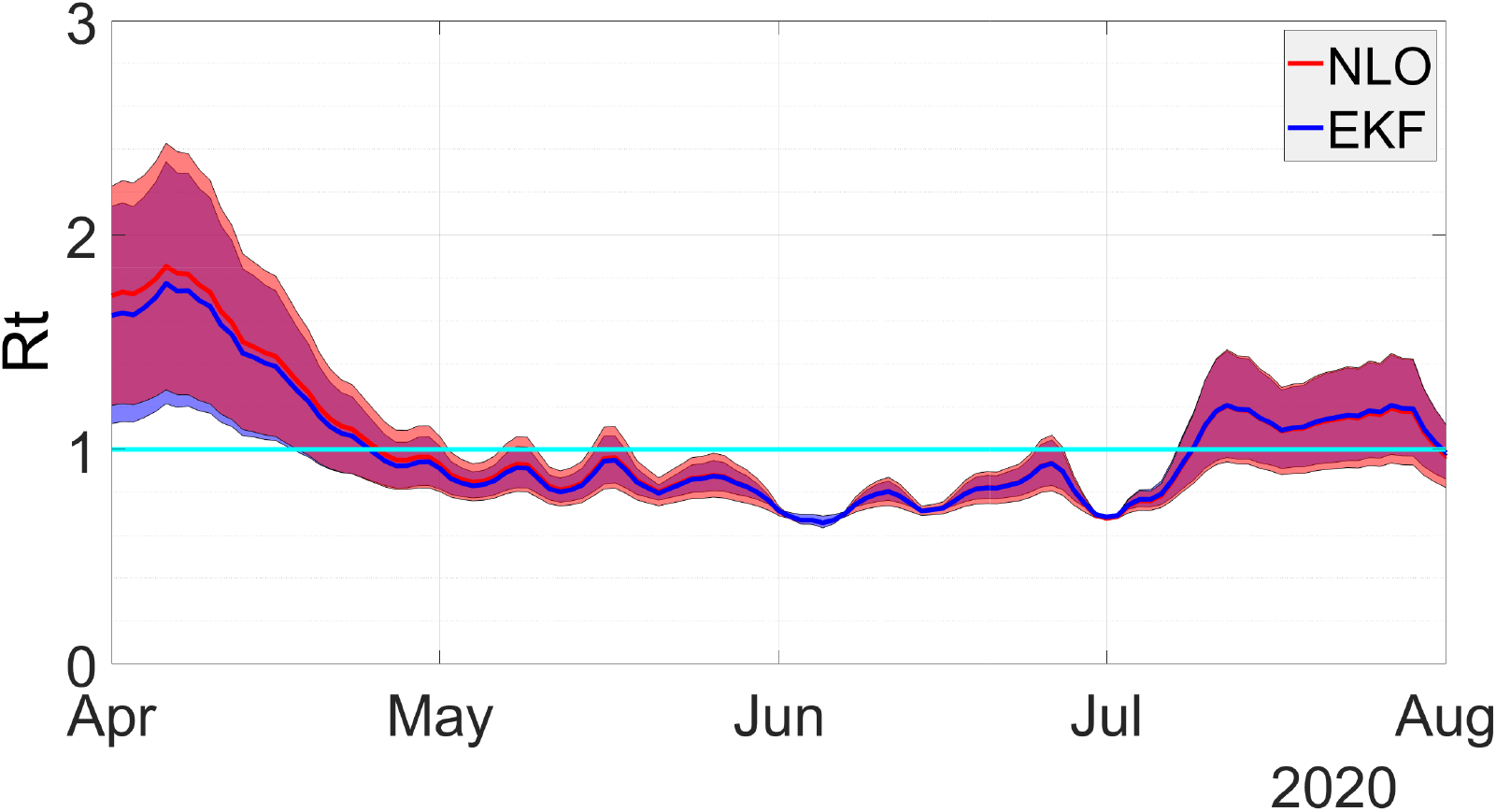
Comparison of the effective reproduction numbers *ℛ*_*t*_ from NLO and EKF. The Confidence Interval (CI), indicated by the band, is a result from uncertainty in the infectious time 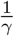

## V. Conclusion

In this paper, we have presented a new approach to estimate the effective reproduction number *ℛ*_*t*_ of infectious diseases. The idea is to inject the discrete-time augmented SIR model with epidemiological data, such as active and removed case. The main challenge is to find a suitable observer gain, since there are no methods to solve the LMI automatically. However, once the observer gain is found, the method is comparable with EKF. The main advantage of using NLO is its stability compared to EKF. Furthermore, it doesn’t require computation of inverse covariance matrices, which makes NLO more efficient.

## Data Availability

Data are available at https://github.com/agusisma/COVIDNLO

